# Delays and disruptions in cervical cancer care services during the COVID 19 pandemic and their impact on survival outcomes – Real world data from India

**DOI:** 10.1101/2025.09.06.25335214

**Authors:** Venkata Krishna Vamsi Gade, Jyoti Sharma, Sougata Maity, Tapesh Bhattacharyya, GY Srinivasa, Dharmendra Kumar Sah, Santam Chakraborty, Bhavana Rai, Sushmita Ghoshal

## Abstract

**Background:** The COVID-19 pandemic led to significant disruptions in cancer diagnosis and treatment in India. This retrospective study with two participating centres aimed to classify and quantify the delays and disruptions in cervical cancer care during the pandemic and evaluate their impact on treatment outcomes.

**Methods:** Records of newly diagnosed patients of cervical cancer registered in between March 2020 and December 2021 were evaluated. A delay group was identified if there was either a gap of more than 3 months from onset of symptoms to histological proof or a gap of more than 6 weeks from diagnosis to start of treatment or a gap of more than 4 consecutive days during Radiotherapy (RT). The cause for the delays and its impact on stage and treatment were identified. Survival curves were plotted using the Kaplan Meir method and differences in survival were analysed using the log rank test.

**Results:** Out of the 418 cervical cancer patients eligible for final analysis registered, 268 (64·1%) were found to have a delay. While a diagnostic delay was noted in 220 (52·6%) patients, a delay in start of treatment was seen in 126 (30·1%) and treatment interruption was seen in 84 (20%) patients. Logistic delays were the most common cause (54%) followed by delays due to COVID 19 infection (7·6%). The delay group had a higher proportion of patients with metastatic disease-Stage IVB (5·9% vs 0·6%) and a lower proportion of patients with early stage disease – Stage I-IIA (7·4 % vs 9·3%) compared to the group without delays(p=0·02). Upward Stage migration was seen in 22% of patients in the Delay group and 11·3% of patients in the No Delay group (p=0·004). Definitive Radiotherapy (RT) followed by Brachytherapy was the most common treatment modality (63·8% of cohort). The proportion of patients receiving external supplementary boost (12·6% vs 9·3%) and Palliative RT (4% vs 0·6%) was higher in the Delay group as compared to the No Delay group (p=<0·001). After a median follow up of 37 months, The 1 year, 2 year and 3 year Overall Survival (OS) was 75%, 58% and 52% in the Delay group and 92%, 87% and 84% in the group without delays respectively (p<0·001).The 1 year, 2 year and 3 year PFS was 65%, 54% and 46% in the Delay group and 91%, 86% and 79% in the group without delays respectively (p<0·001).

**Conclusion:** Delays and disruptions in cervical cancer care during the COVID-19 pandemic led to tumour upstaging and worsened survival outcomes. This highlights the urgent need for developing strategies aimed at optimal resource utilisation and minimizing delays in the event of a future pandemic.

**Research in Context:** *WHAT IS ALREADY KNOWN ON THIS TOPIC:* The COVID-19 pandemic severely disrupted cancer care, worsening delays in diagnosis and treatment, particularly in low- and middle-income countries like India. Cervical cancer patients faced increased risks due to interruptions in radiotherapy and diagnostic delays. It is well documented that prolongation of overall treatment time can adversely affect treatment outcomes in patients with cervical cancer .

*WHAT THIS STUDY ADDS:* To our knowledge, this retrospective cohort study from two institutions is the first study that reports the real-world impact of the COVID 19 pandemic on cervical cancer survival outcomes in India. In this study we describe and stratify the various kinds of delay – namely diagnostic delay, delay in initiation of treatment and delay during treatment. We demonstrate increased incidence of upward stage migration in patients with delays. The overall survival and progression free survival was significantly worse in the group with delays as compared to the group without delays. We also show that most of the delays are attributable to logistic reasons rather than COVID 19 infection. Delays caused by COVID 19 did not significantly influence survival outcomes.

*HOW THIS STUDY MIGHT AFFECT RESEARCH, PRACTICE, OR POLICY:* The findings of this study seem to suggest that COVID 19 based nationwide lockdowns, inadvertently led to significant disruptions in cervical cancer care, resulting in poorer patient outcomes. This study highlights the urgent need for developing strategies aimed at optimal resource utilisation and minimizing delays in the event of a future pandemic.

## Introduction

The COVID 19 infection caused by the novel coronavirus (SARS-CoV-2) was declared a pandemic by the World health organisation (WHO) on 11^th^ March 2020^1^ . In response, various countries enacted lockdowns that restricted the movement of people across national, state and district borders in an effort to the control the rise in the number of cases ^2^. Indian authorities imposed a nationwide lockdown on March 25^th^, 2020 with restrictions on movement titrated based on the case load. It is considered that the first wave of the pandemic in India occurred between March 2020 and January 2021 whereas the more severe second wave occurred between March 2021 and December 2021 ^3–5^.

The COVID 19 pandemic disrupted cancer care services globally. Patients suffering from cancer were at a greater risk of hospitalisation and death after contracting the COVID-19 infection ^6^. As of the end of 2021, WHO reported that upto 50% of the cancer screening and treatment programs were disrupted amongst all reporting countries ^7^. The effects were disproportionately felt in Low Middle income countries like India where the pandemic exacerbated existing disparities in access to cancer care. A large cohort study from India revealed that there was a 38% reduction in the pathological diagnostic tests and 23% reduction in the number of patients accessing radiotherapy during the pandemic ^8^.

During the treatment of cervical cancer, the prolongation of the overall treatment time due to interruptions in radiotherapy adversely affects survival and local control ^9^. Diagnostic delays and delays in treatment initiation lead to upward stage migration which also worsens outcomes ^10, 11^. A study from a South Indian centre reported that prior to the pandemic, the median time between detection of symptoms and diagnosis of cervical cancer was 80 days and that between detection of symptoms and start of treatment was 123 days ^12^. The current study was designed to qualify and quantify the diagnostic and treatment delays in cervical cancer treatment and their effect on survival during the COVID 19 pandemic at two Indian centres.

## Materials and methods

This was an observational retrospective cohort study with two participating Indian tertiary cancer centres. The center located in Northern India (C1) was a public funded superspeciality hospital with a regional cancer center. The center in Eastern India (C2) was a charitable trust funded dedicated cancer hospital. . Institute ethical committee clearances were obtained from both the centres. Deidentified data was shared from C2 to C1 for the analysis. The primary objective of the study was to quantify and qualify the delays in diagnosis and treatment of patients with cervical cancer treated during the COVID pandemic. The secondary objectives were to analyse the impact of delays on Overall survival (OS) and Progression free survival (PFS)

Records of patients with newly diagnosed cervical cancer registered between March 2020 and December 2021 were evaluated. While data of all treated cases were included from C1, data patients treated with curative intent definitive radiotherapy were included from C2. Thus the dataset from C2 did not include patients receiving adjuvant radiotherapy after surgery or palliative radiotherapy. . Various data variables were recorded by accessing the physical (C1) or electronic (C2) health records. Patient identifiers such as name and address were masked for the purpose of the study. Patient-related variables collected included Age, performance status, pre-treatment renal function and hematological parameters. Disease-related parameters included Stage at diagnosis, Stage at start of therapy, presence of lymph nodes and presence of Hydroureteronephrosis (HDUN). The treatment related details included information regarding the intent of treatment, surgery, radiotherapy, brachytherapy (BT) and chemotherapy.

A delay group was identified if there was a gap of more than 3 months between onset of symptoms and histological diagnosis (Diagnostic delay), or a gap of more than 6 weeks from the date of diagnosis to start of treatment (Delay in treatment initiation) or a gap of more than 4 consecutive days during chemoradiation (CRT) (Delay during radiotherapy). Two separate groups of patients – the “Delay Group” and the “No Delay Group” were identified.

### Measures to address bias and loss to follow up

To minimize selection and interpretation bias – two investigators from each centre verified every entry with the digital or electronic health record. Patients that were registered at C1 but were not treated at that centre were categorised as Registered Only. Patients designated as Registered Only were contacted telephonically and requested to visit the treating centre for the purpose of recording the details of treatment taken elsewhere. Patients that did not report to the centre after treatment completion were also telephonically contacted and urged to visit the treating centre. In cases where a patient was unable to attend the centre, the patient was encouraged to follow up locally and provide the records at the next possible visit to the centre. Patients that did not report for follow up and with whom telephonic contact was unsuccessful were designated as Lost to Follow up (LFU). Patients that have been categorised as Registered only were included in the analysis if treatment records were retrievable. Patients that were categorised as both Registered only and LFU were removed from the analysis.

### Statistical analysis

Study related variables were entered into SPSS v25 (SPSS –Statistical package for social sciences – IBM). Two groups were delineated as outlined above – “DELAY group” and “NO DELAY group”. Descriptive data was generated for variables in the study. Independent samples T-test was used to analyse normally distributed continuous data and Mann Whitney U test was used to analyse non normal continuous data. Chisquared and Fischers exact test was used to compare categorical variables in both groups.

Overall survival (OS) was calculated from date of diagnosis to date of death due to any cause. Progression free survival (PFS) was calculated from date of diagnosis to date of progression or death. Survival analysis was done by Kaplan Meir method and survival outcomes were compared using the log-rank test. Median follow up duration was calculated by the reverse Kaplan Meir Method. Univariate and Multivariate analysis was done with the Cox Proportional hazards method. Multivariate models were run separately for patients treated with curative chemoradiotherapy and those treated with palliative intent .As this was an observational retrospective study where the electronic and paper records of patients were used for collecting data, there was no direct interaction with subjects of the study.

## RESULTS

### Baseline features

418 patients met the inclusion criteria and were eligible for final analysis (Figure 1). While 268 patients belonged to the Delay Group, 150 patients belonged to the No Delay Group. 331 patients belonged to C1 and 87 patients belonged to C2. While 74·3 % of patients registered at C1 had a delay, only 25·3% of patients registered at C2 had a delay. The mean age was similar in both the groups. In the entire cohort, 8·1% of patients belonged to early stage (I-IIA), 87·7% of patients belonged to Locally Advanced Stage (IIB-IVA) and 4% had metastatic disease. The delay group had a higher proportion of patients with metastatic disease and a lower proportion of patients with early-stage disease. Stage migration (disease progression in between diagnostic scans and simulation scans) was seen in 22% of patients in the Delay group and 11·3% of patients in the No Delay group. Squamous cell Carcinoma was the most common histological type (92·8%) followed by Adenocarcinoma (4%) and other histology (Poorly differentiated Carcinoma, Clear cell Carcinoma, Neuroendocrine Carcinoma). 86·3% of patients in the cohort were treated with curative intent and 7·1% of patients were treated with palliative intent. Twenty-seven patients (6·4%) (all in the Delay Group) did not undergo the prescribed treatment (Table 1). A centre wise break-up of the baseline characteristics in provided in Appendix Table 1.

**Figure 1.**
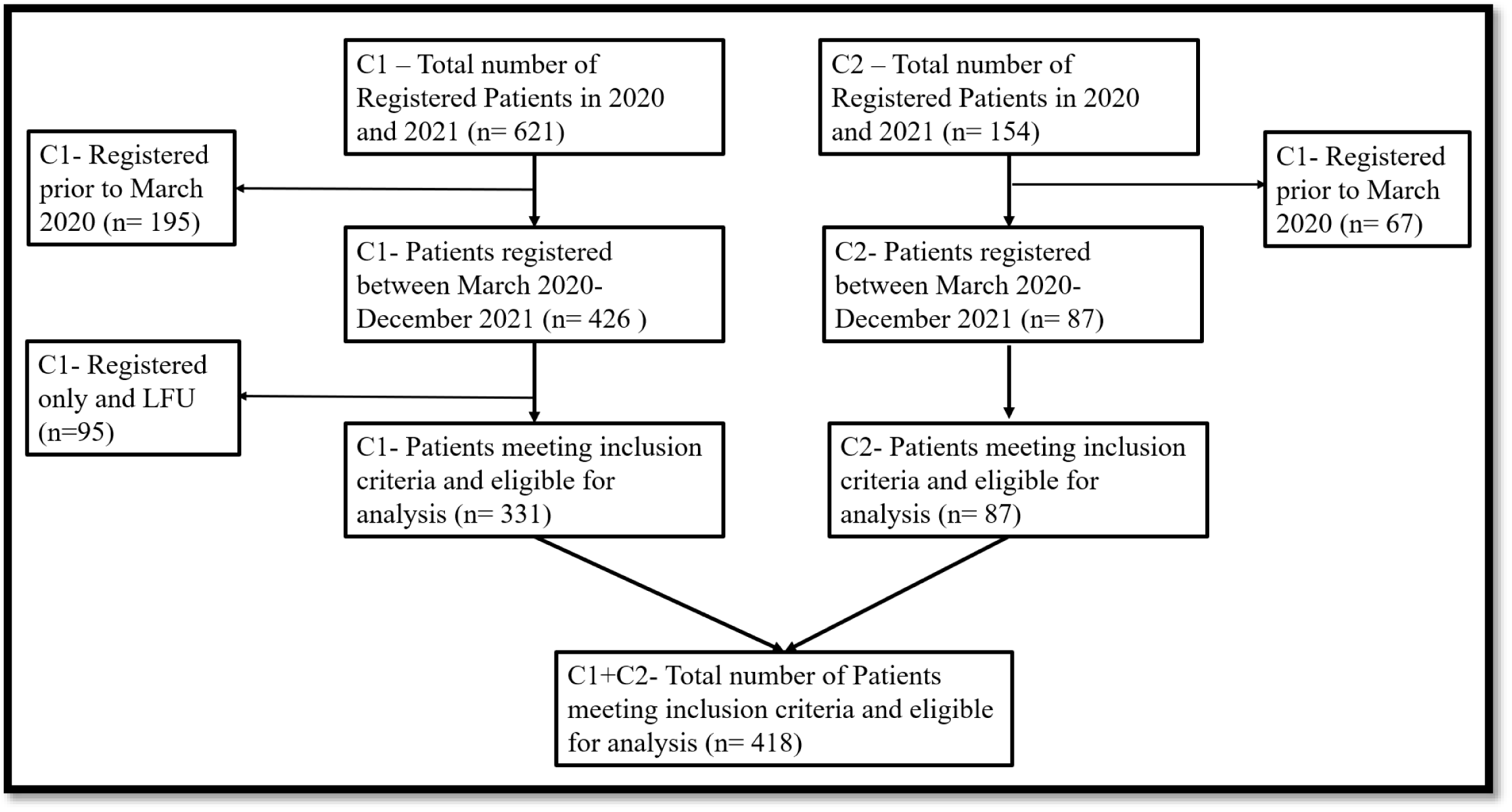
Description of study population. (C1-Centre 1; C2-Centre 2, LFU – lost to follow up)

**Table 1.** Clinical features of the study population.

| Feature | Entire cohort<br>(n= 418) | Delay Group<br>(n=268) | No Delay<br>Group<br>(n=150) | p Value |
| --- | --- | --- | --- | --- |
| Age (in years)<br>Mean +/- SD | 54·09<br>(±10·8) | 54·06<br>(±10·8) | 54·14<br>(±10·9) | 0·943 |
| Stage |  |  |  |  |
| I-IIA | 34 (8·1%) | 20 (7·4%) | 14 (9·3%) | **0·02** |
| IIB-IVA | 367 (87·7%) | 232 (86·5%) | 135 (90%) |  |
| IVB | 17 (4%) | 16 (5·9%) | 1 (0·6%) |  |
| Stage migration | 76 (18·1%) | 59 (22%) | 17 (11·3%) | **0·004** |
| Histology |  |  |  |  |
| Squamous cell | 388 (92·8%) | 244 (91%) | 144 (96%) |  |
| Carcinoma | 17 (4%) | 14 (5·2%) | 3 (2%) | 0·479 |
| Adenocarcinoma | 13 (3·1%) | 10 (3·7%) | 4 (2%) |  |
| Others |  |  |  |  |
| Treatment intent |  |  |  |  |
| Curative | 361 (86·3%) | 216 (80·5%) | 145 |  |
| Palliative | 30 (7·1%) | 25 (9·3%) | (96·6%) | **<0·001** |
| Not treated | 27 (6·4 %) | 27 (10 %) | 5 (3·3%) |  |
|  |  |  | 0 (0%) |  |

### Treatment details and Patterns of failure

Definitive Radiotherapy (RT) followed by Brachytherapy was the most common treatment modality (63·8% of cohort). 11.4% of patients received external supplementary boost. This was followed by Surgery and Adjuvant RT in 7·1%, Palliative RT in 2·8% and Palliative Chemotherapy in 2 patients (0·4%). The proportion of patients receiving external supplementary boost and Palliative RT was higher in the Delay group as compared to the No Delay group. The median follow-up duration was 29 months (IQR -11-39.2 months). 12·6% of patients in the No delay group and 33·3% of patients in the Delay group had disease progression or treatment failure on follow up. In the entire cohort, forty-eight (11·4%) patients had both Local and Regional recurrence, and Thirty-eight (9%) patients had Local, regional and metastatic recurrence (Table 2). A centre wise break up of the treatment related details and patterns of spread is provided in Appendix Table 2.

**Table 2.**
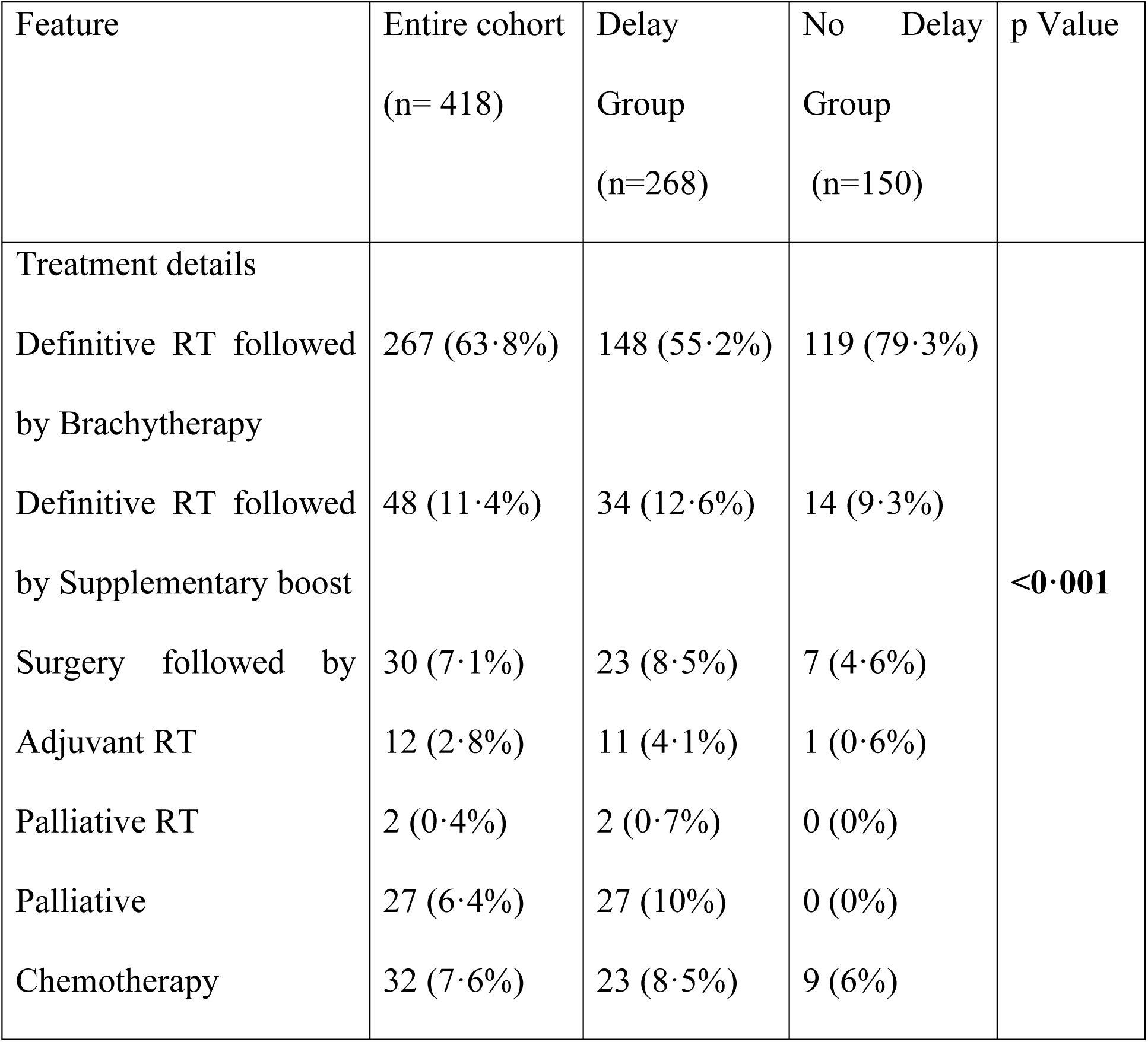
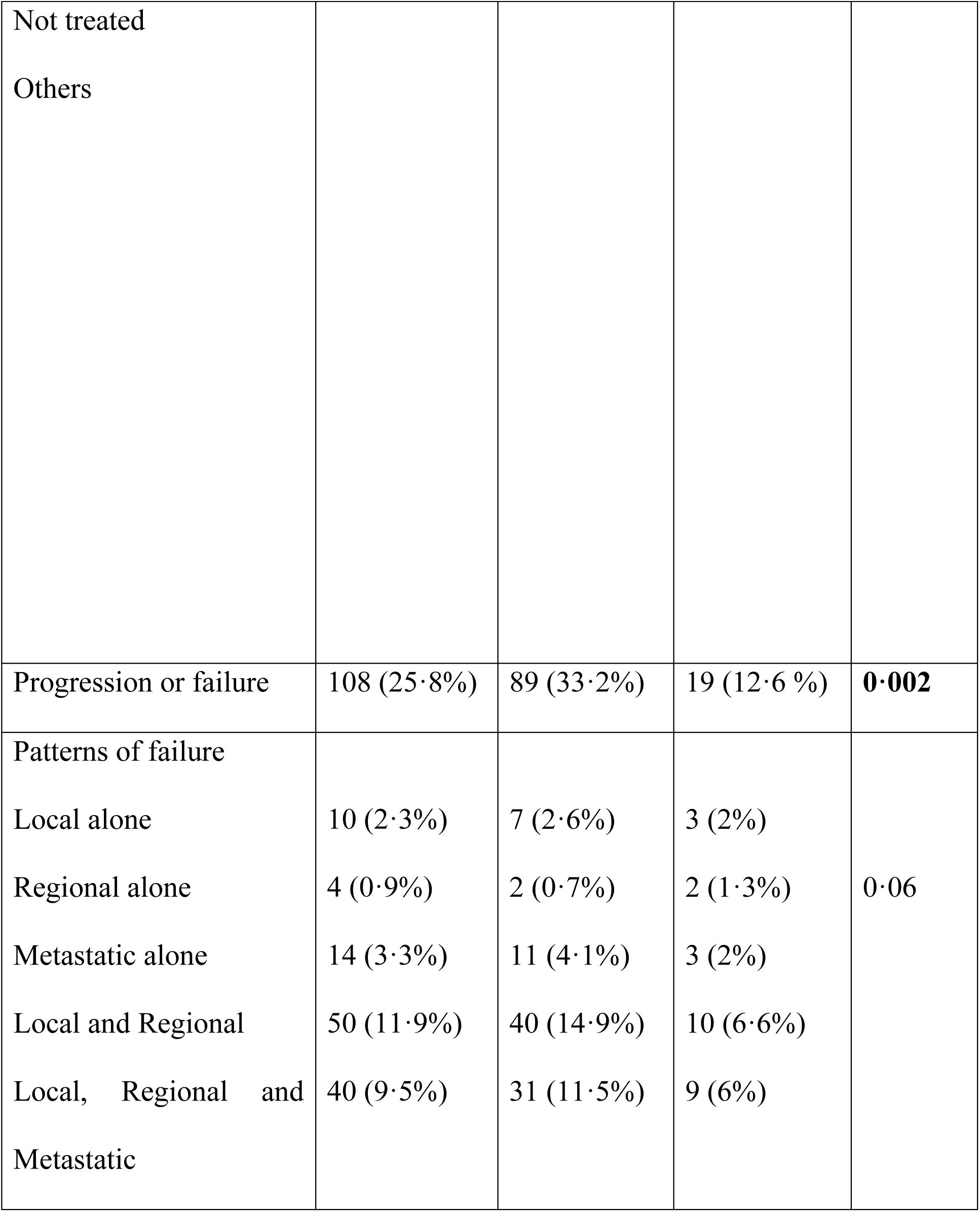
Treatment related details and patterns of spread (Note – Others in the treatment column includes those that received EBRT alone but no brachytherapy, surgery alone without adjuvant radiotherapy and those offered best supportive care)

### Types of Delay and its features

A diagnostic delay was noted in 220 patients (52·6%). A delay in start of treatment was seen in 126 (30·1%) patients. Treatment interruption was seen in eighty-four (20%) patients. (Figure 2). Ninety patients had only a delay in diagnosis. While Seventy (16·7%) patients had a delay in both diagnosis and start of treatment, Thirty-six (8·6%) patients had both a delay in diagnosis and a delay during RT. Twenty-five (5·9%) patients had a delay only in the start of treatment. While Seventeen (4%) patients had a delay only during RT, Eight patients had a delay in start of treatment and during RT. While 226 (54%) patients had delay due to logistic issues, thirty-two patients (7·6%) had delay due to COVID 19 infection

**Figure 2.**
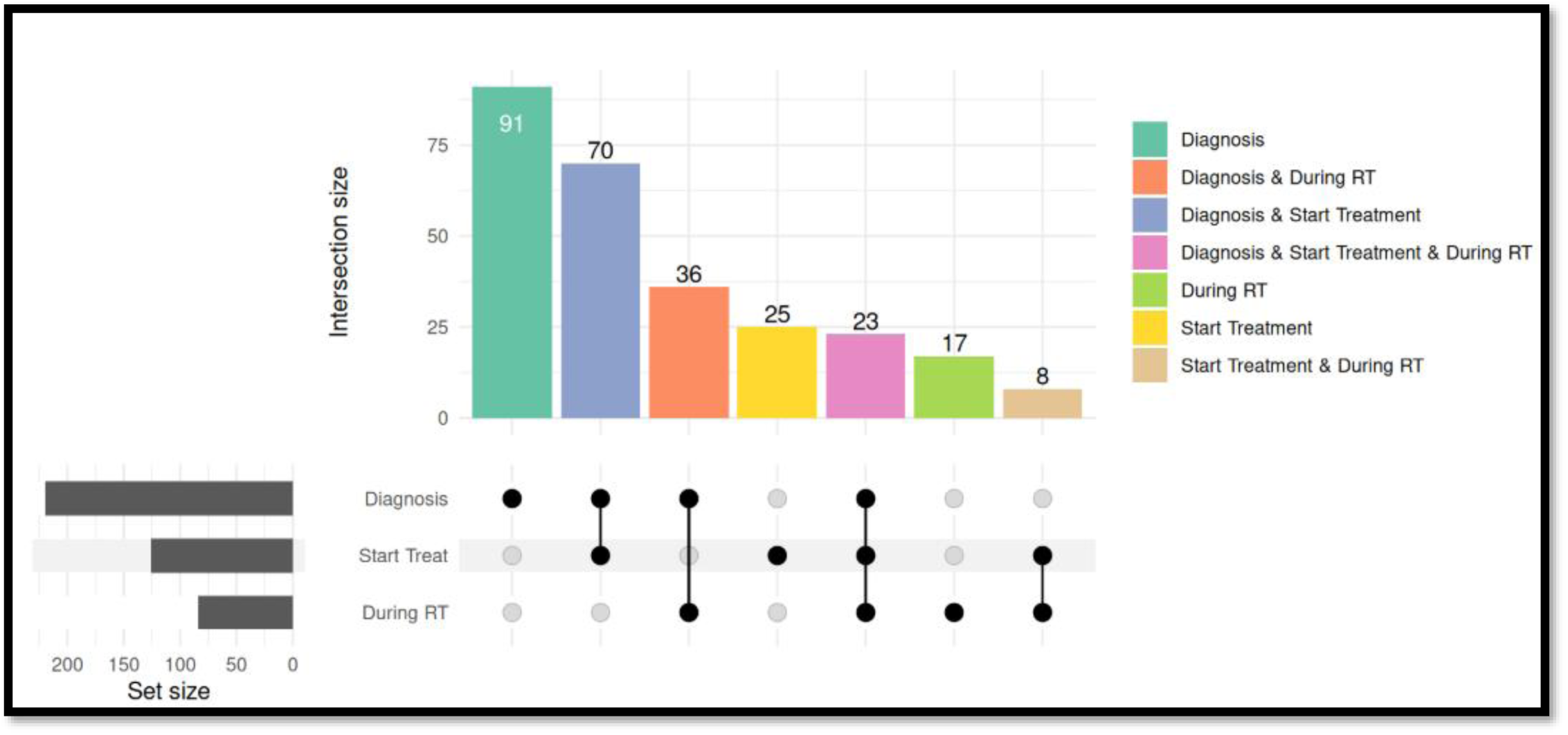
Upset plot illustrating the various types of delay in cervical cancer care in the study population (n=418).

### Survival analysis

The median follow-up duration was 37 months (95% CI – 35·8 -38·1 months). The Median OS was 38 months (95% CI – 28·7 – 47·2 months) in the Delay group and Not Reached in the No delay group. The 1 year, 2 year and 3 year Overall Survival (OS) was 75%, 58% and 52% in the Delay group and 92%, 87% and 84% in the group without delays respectively (p **<0**·**001**). The Median Progression Free Survival (PFS) was 31 months (95% CI – 18.7 – 43.2 months) in the Delay group and Not Reached in the No delay group. The 1 year, 2 year and 3 year PFS was 65%, 54% and 46% in the Delay group and 91%, 86% and 79% in the group without delays respectively (p<0·001). (Figure 3 and 4). Amongst patients treated with curative intent, the 1yr, 2 yr and 3 yr OS was 83%, 67% and 61% in the Delay group and was 93%, 89% and 86% in the group without delays (Appendix Table 3)

**Figure 3.**
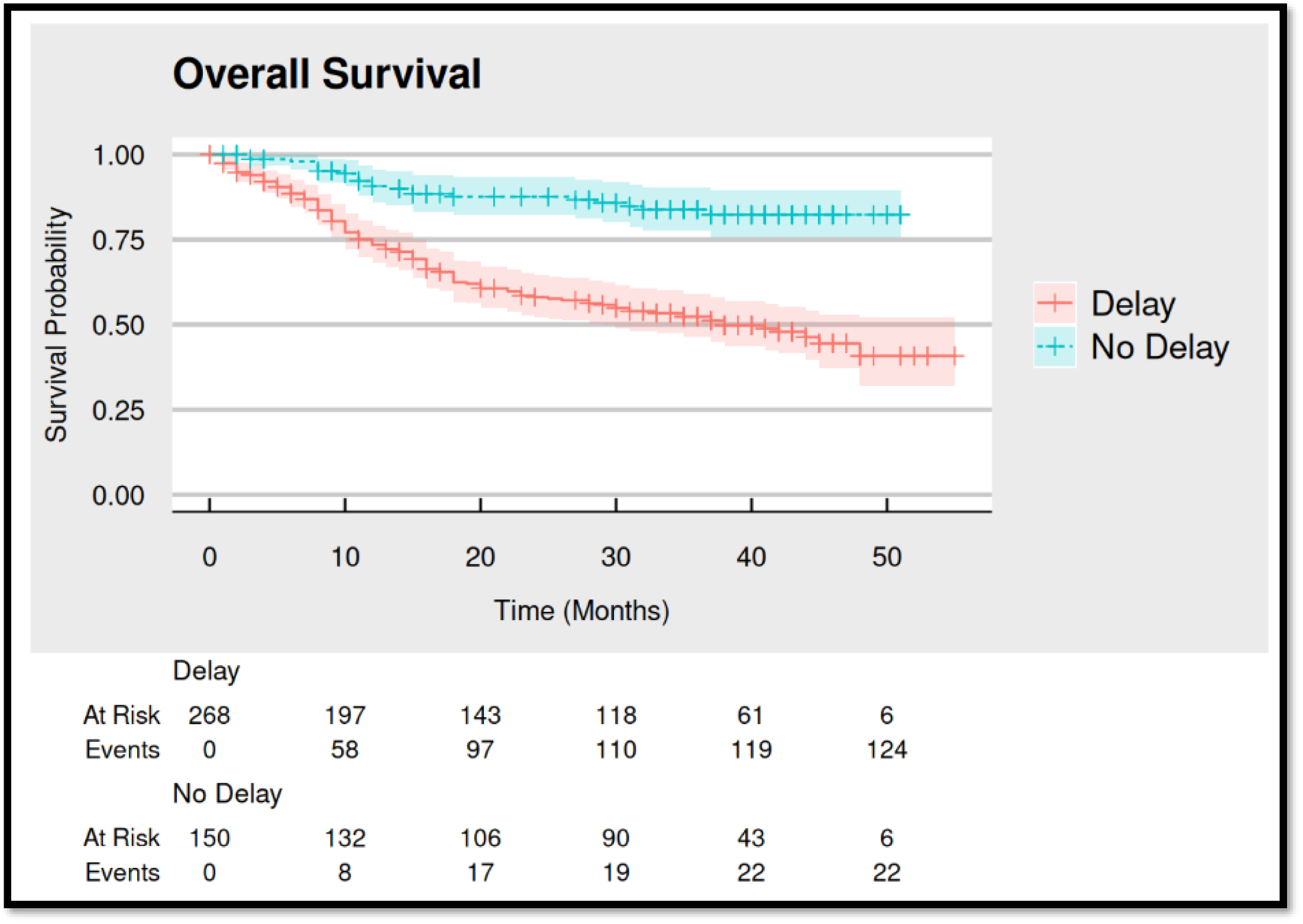
Kaplan Meir curves for the Overall survival (OS) for both groups of the study population

**Figure 4.**
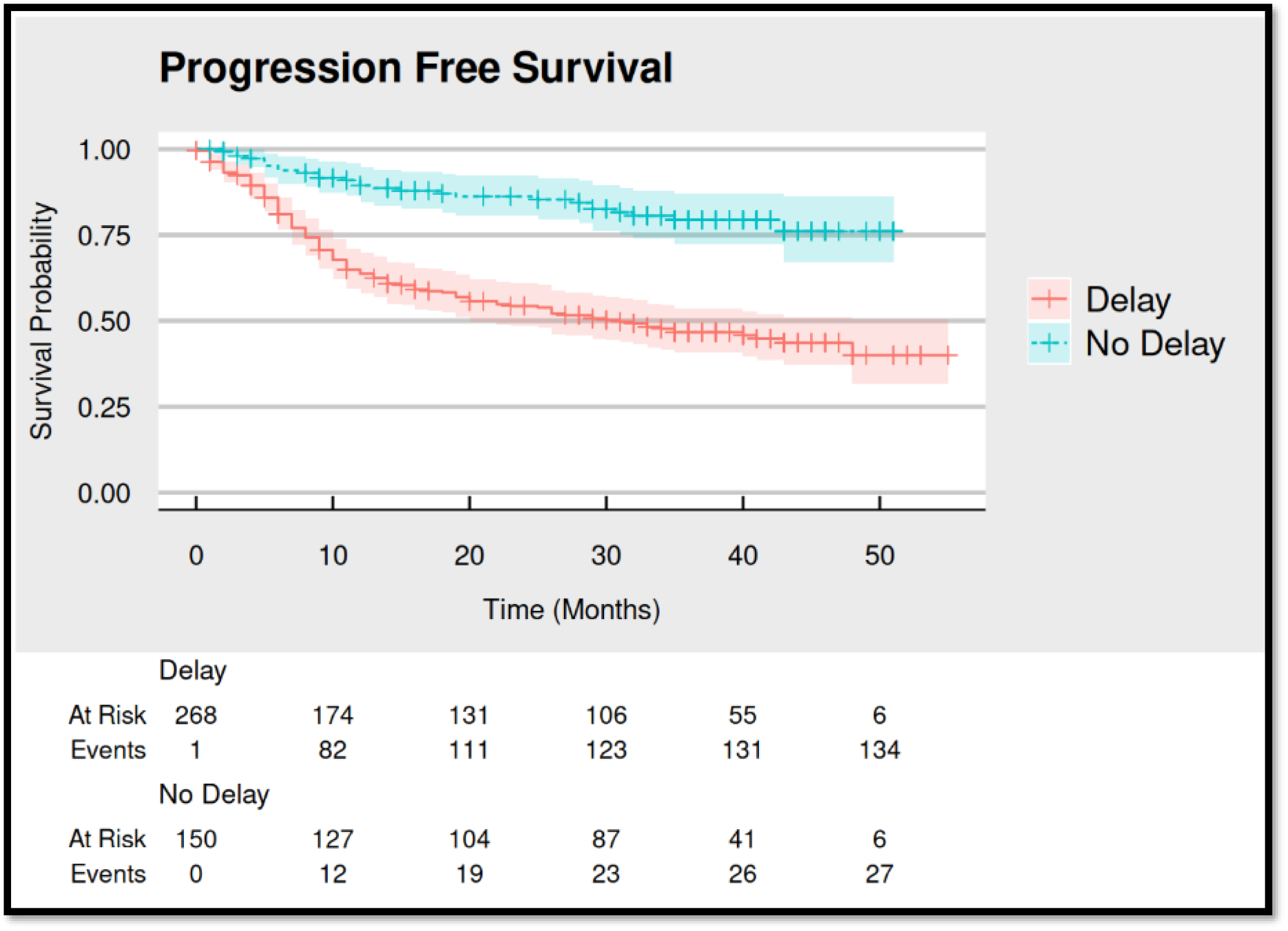
Kaplan Meir curves for the Progression free survival (OS) for both groups of the study population

Univariate analysis revealed that pre-treatment Hemoglobin, Pre-treatment creatinine, HDUN, administration of concurrent chemotherapy were significant predictors of OS. Similarly, those that had a delay in diagnosis, delay in starting treatment and those that had delay due to logistic issues seemed to have adverse OS outcomes (Table 3). Amongst patients treated with curative chemoradiotherapy followed by brachytherapy, log relative hazard plots (nomogram) from multivariate analysis show that increase in duration of symptoms trended towards worse survival outcomes with an inflection region seen after 300 days. Pre-treatment Hemoglobin of more 10 g/dL was associated with better OS trends (Figure 5 and Table 4). A second multivariate model was performed where instead of studying delay as a numerical variable, the presence of delay was studied as a categorical variable. In that analysis, pre-treatment Hemoglobin and presence of delay affected OS trends (Appendix Figure 1, Table 4). Similar trends were observed in PFS outcomes (Appendix Tables 5,6, Appendix Figures 2,3). Amongst patients treated with palliative intent, increase pre-treatment creatinine and increase in duration of symptoms predicted worse survival outcomes (Appendix table 7, Appendix Figure 4)

**Figure 5.**
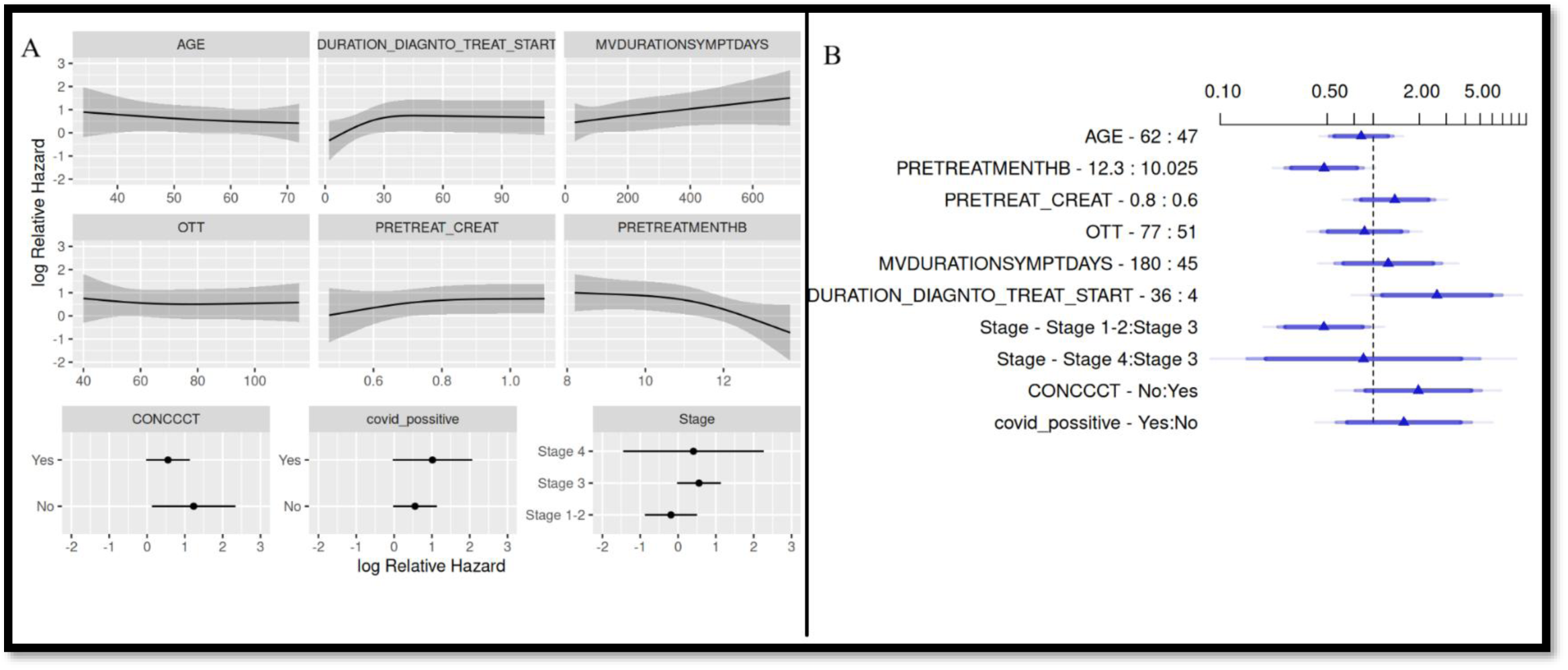
(A) - Log relative hazard plots for multivariate analysis of various factors affecting the OS in patients treated with curative chemoradiotherapy. (B)-Cox model Forest plot for multivariate analysis of various factors affecting the OS patients treated with curative chemoradiotherapy. (OTT – Overall treatment time, MVDURATIONSYMPTOMDAYS – Duration of symptoms in days, CONCCT – Concurrent chemo given or not)

**Table 3.**
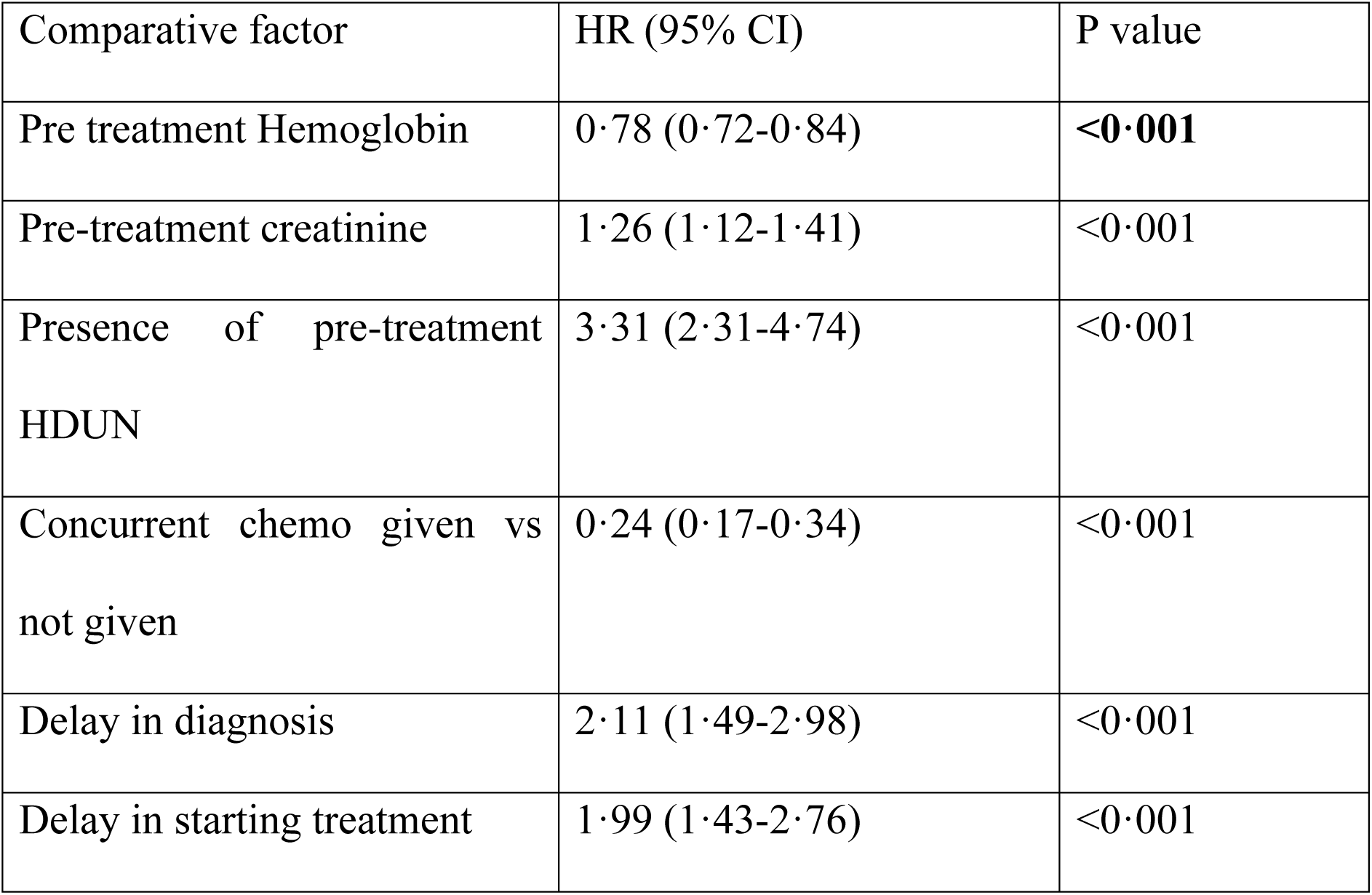
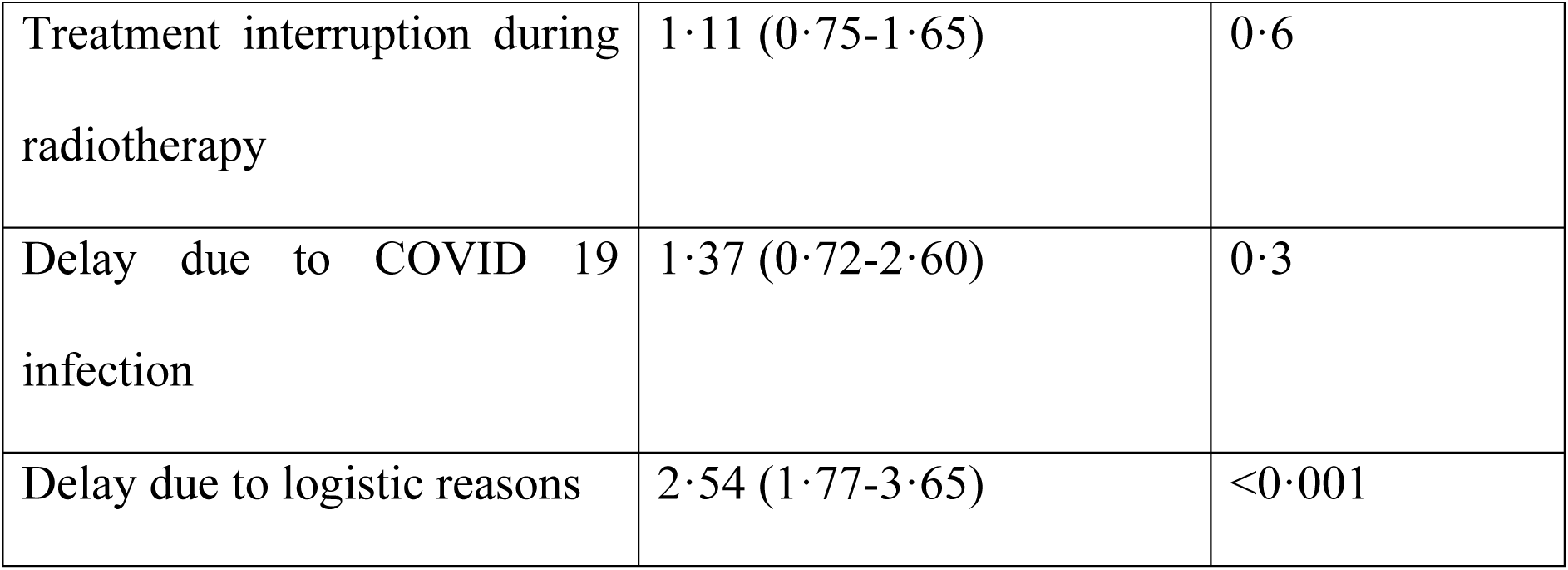
Univariate Cox Proportional Hazard analysis of factors affecting the OS.

| Comparative factor | HR (95% CI) | P value |
| --- | --- | --- |
| Pre treatment Hemoglobin | 0·78 (0·72-0·84) | **<0·001** |
| Pre-treatment creatinine | 1·26 (1·12-1·41) | <0·001 |
| Presence of pre-treatment HDUN | 3·31 (2·31-4·74) | <0·001 |
| Concurrent chemo given vs not given | 0·24 (0·17-0·34) | <0·001 |
| Delay in diagnosis | 2·11 (1·49-2·98) | <0·001 |
| Delay in starting treatment | 1·99 (1·43-2·76) | <0·001 |
| Treatment interruption during radiotherapy | 1·11 (0·75-1·65) | 0·6 |
| Delay due to COVID 19 infection | 1·37 (0·72-2·60) | 0·3 |
| Delay due to logistic reasons | 2·54 (1·77-3·65) | <0·001 |

**Table 4.**
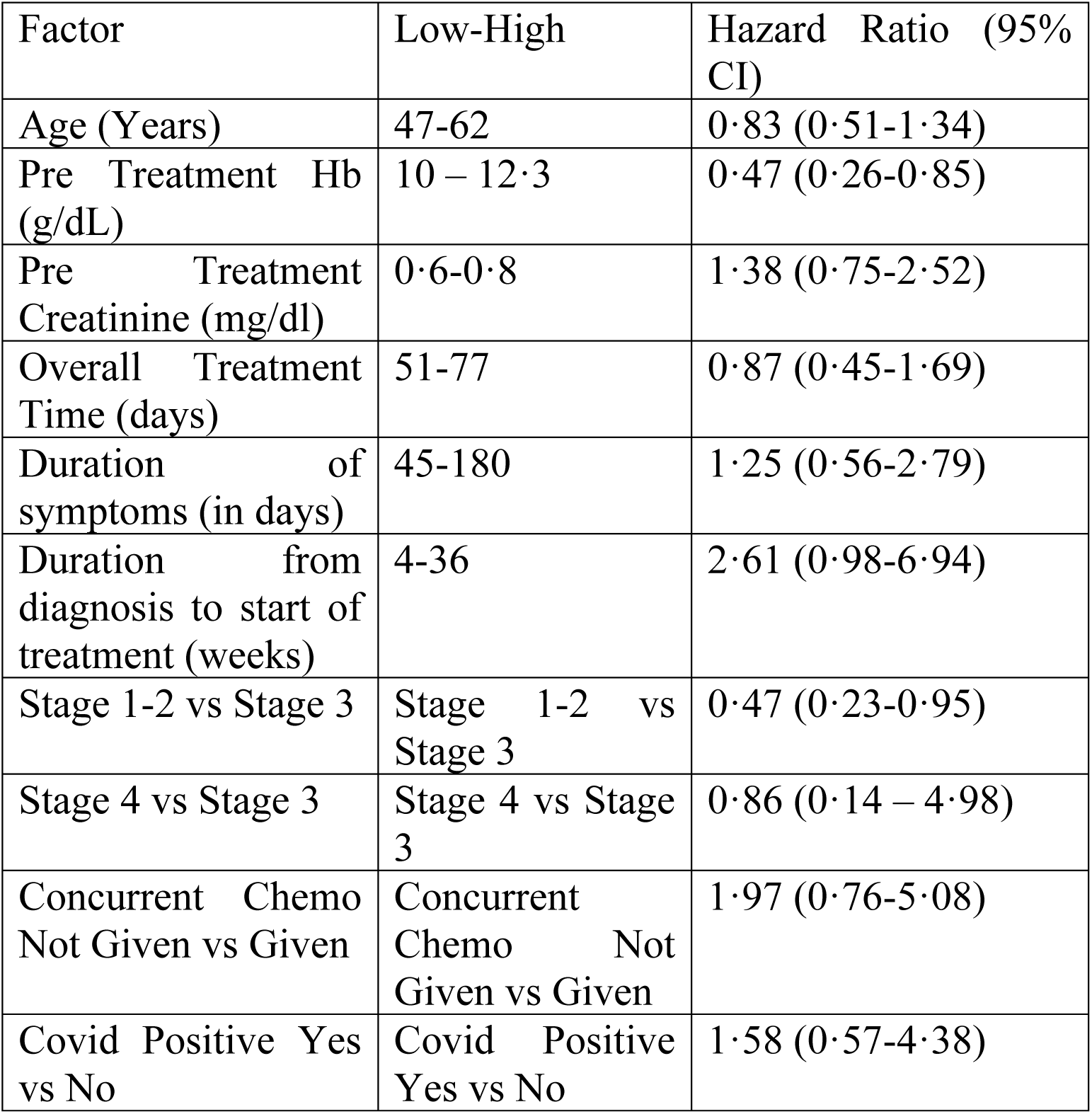
Multivariate analysis of various factors affecting the OS patients treated with curative chemoradiotherapy.

| Factor | Low-High | Hazard Ratio (95% CI) |
| --- | --- | --- |
| Age (Years) | 47-62 | 0·83 (0·51-1·34) |
| Pre Treatment Hb (g/dL) | 10 – 12·3 | 0·47 (0·26-0·85) |
| Pre Treatment Creatinine (mg/dl) | 0·6-0·8 | 1·38 (0·75-2·52) |
| Overall Treatment Time (days) | 51-77 | 0·87 (0·45-1·69) |
| Duration of symptoms (in days) | 45-180 | 1·25 (0·56-2·79) |
| Duration from diagnosis to start of treatment (weeks) | 4-36 | 2·61 (0·98-6·94) |
| Stage 1-2 vs Stage 3 | Stage 1-2 vs Stage 3 | 0·47 (0·23-0·95) |
| Stage 4 vs Stage 3 | Stage 4 vs Stage 3 | 0·86 (0·14 – 4·98) |
| Concurrent Chemo Not Given vs Given | Concurrent Chemo Not Given vs Given | 1·97 (0·76-5·08) |
| Covid Positive Yes<br>vs No | Covid Positive<br>Yes vs No | 1·58 (0·57-4·38) |

## Discussion

This retrospective cohort study from two centers in India was designed to evaluate the impact of COVID 19 related delays and disruptions in cervical cancer on survival outcomes. Out of the 418 patients evaluated, 64·1% were found to have a delay. Patients in the delay group had a higher proportion of patients with metastatic disease and upward stage migration as compared to the no delay group. After a median follow up of 29 months, the OS and PFS outcomes were significantly worse in patients belonging to the Delay group as compared to the group without delays. Anemia, Pretreatment Renal dysfunction and advanced stage were significant predictors of adverse survival outcomes.

According to data from the WHO, about 7 million people have died as a result of the COVID 19 infection worldwide with about five hundred thousand deaths occurring in India. Various epidemiological studies estimate the excess mortality during the pandemic period to be around 18 million ^13–17^. This includes excess deaths that have directly or indirectly occurred as a result of the COVID pandemic. The COVID pandemic led to a sustained disruption in cancer care programs around the world. The Veterans Affairs group reported that new cancer diagnoses decreased by 13 % during the pandemic ^18^ . A study from the UK revealed a 70% decline in urgent referrals and 31% decline in chemotherapy visits in the early months of the pandemic ^19^. About 50% of the cancer care programs were disrupted by the end of 2021 – an effect that was disproportionately felt in low and low middle income countries ^7^. Indian authorities imposed a nationwide lockdown on March 25^th^, 2020 where the extent of restrictions on movement across state and district lines were titrated based on the case load ^2, 3^.

In the current study, while 74·3 % of patients registered at C1 had a delay, only 25·3% of patients registered at C2 had a delay. C1 is large tertiary care Institute of national importance under the Union Ministry of Health and Family Welfare, Government of India. During the pandemic, C1 functioned as an apex referral centre for COVID care for patients from four north Indian states. Large sections of the hospital were set aside for COVID care. Significant numbers of hospital employees from various departments were engaged in COVID care. The Department of Radiotherapy at C1 continued to function during the pandemic with limited OPD capacity and without inpatient facilities. Telemedicine services were used to supplement the OPD by attending to patients on follow that did not have any symptoms and to streamline OPD appointments. The brachytherapy Operation theatre (OT) and chemotherapy day care functioned with reduced capacity. Due to the lack of inpatient facilities and limited OT slots, about 46 patients of cervical cancer at C1 with significant residual disease after Definitive radiotherapy could not receive advanced brachytherapy application and instead received External supplementary boost. About 22 patients did not receive concurrent chemotherapy due to these logistic issues. In contrast, C2 is a dedicated cancer hospital run by a Private Charitable trust. During the duration of the pandemic, C2 had retained access to OTs, Inpatient facilities and chemotherapy day-care facilities as they were not a designated COVID care centre. C2 however, provided Covid treatment to patients who acquired the infection during the cancer treatment.

A Population based study from Romania found that in the initial months of the pandemic-there was a 75·5% reduction in the volume of diagnostic tests performed for cervical cancer. While the number of newly diagnosed cases reduced by 45% in the 24 months of the pandemic, they reported a 17% increase in the number of Stage III-IV cancers ^20^ .A cohort study from 41 cancer centers in India showed that – during the first three months of the pandemic, there was a 54% reduction in the new cases registered, 37% reduction in patients accessing chemotherapy and 23% reduction in patients accessing radiotherapy ^8^. An audit from RCC Trivendrum revealed that there was a 36% reduction in the number of newly diagnosed breast cancer cases in 2020 as compared to 2019 with geographical variation in the state of Kerala. However, it did not seem to impact diagnosis and treatment of breast cancer patients ^21^. An audit on breast cancer patients from our institute showed that about 15·5% experienced a COVID 19 related delay in diagnosis or delay in start of treatment. Amongst those with a delay, 45·7% had cancer upstaging or disease progression ^22^. In the current study, 64·1% of the cohort was found to have a delay with 22% of patients in the Delay group having upward stage migration in between their diagnostic and simulation scans.

Studies from western countries appear to suggest that pandemic induced lockdowns disrupted cervical cancer screening and vaccination programs leading to a potential increase in the number of cases ^20, 23^ .India being Low middle income country (LMIC) already has a high burden of cervical cancer with 127,526 new cases in 2022 ^24^. Here, COVID related delays have the potential to impact survival rather than incidence due to delays in diagnosis and treatment initiation. A model study from India estimated a 2·5-3·8% lifetime increase in deaths due to cervical cancer due to the delays caused during the pandemic ^25^. Prolongation of the overall treatment time in the treatment of cervical cancer adversely affects survival and local control outcomes^9^. A study conducted in Brazil from 2012-2017 showed that delays in cervical treatment conclusion adversely affected 36 month death and survival risk. However, delays in treatment initiation did not correlate with worse survival^10^. A Meta-analysis by Shimels et al of eleven studies with about 50,000 patients found that a 4-week delay in treatment was associated with a 1·27 times higher rate of mortality at 5 years follow up ^11^ .In the current study, we report significantly worse OS and PFS in patients of cervical cancer with diagnostic or treatment delays.

Only 7·6% of patients in the cohort had a delay in diagnosis or treatment due to COVID 19 infection. In the current study, univariate analysis shows that delays in diagnosis and treatment caused due to COVID infection did not adversely affect survival. Most the delays in this current study have been attributed to logistic reasons. Patients’ fear of contracting the COVID-infection in combination with lockdown related restrictions to movement hampered timely diagnosis and initiation of treatment ^26^. The findings of this study seem to suggest that COVID 19 based nationwide lockdowns, inadvertently led to significant disruptions in cervical cancer care, resulting in poorer patient outcomes. While the intent was public safety, the restrictions imposed, lack of tailored guidance, and diversion of resources to COVID care systematically compromised the continuum of cancer care—from diagnosis to treatment. These delays manifested at every stage: diagnosis, initiation of therapy, and interruption of radiotherapy, leading to tumour upstaging and reduced survival rates.

There is an urgent need to formulate site wise cancer care guidelines in the event of another pandemic. Such guidance should be tailored to LMICs and should be part of an overall pandemic preparedness plan. Possible strategies to improve cancer services during a pandemic should focus on both diagnostic and treatment related delays. An observational study analysing the case load from 27 countries found that while lockdowns reduced daily caseloads they did not have much effect on overall prevalence and mortality due to COVID 19 infection ^27^. Diagnostic and treatment related delays to many curable medical conditions could partly explain the excess mortality during this period ^15, 16, 19^. In the event of future pandemic, lawmakers could take these factors into consideration before imposing significant restrictions to movement ^2, 3^. Other measures to mitigate diagnostic delays could include increased awareness about the signs and symptoms of cervical cancer. Improved access to screening and HPV vaccination programs could reduce the overall burden of Cervical cancer which could lead to more effective utilisation of limited resources in the event of a pandemic ^28^. Telemedicine services could be used to streamline outpatient visits by identifying patients with symptoms that need a priority visit for diagnosis or start of treatment ^29^.

Some authors suggest the use of hypofractionated radiotherapy (HFRT) schedules to reduce treatment duration. A systematic review concluded that while HFRT of 40Gy in 16 fractions is comparable to conventional fractionation in terms of outcomes and safety, the usage of >3Gy/ fraction appears to be associated with increased acute and late toxicity ^30, 31^. The use of Abbreviated brachytherapy schedules with multiple fractions treated with a single application is another promising strategy that can optimally utilise limited OT slots in the event of a pandemic. A retrospective study of 64 patients of locally advanced cervical cancer treated with brachytherapy schedules of 3-5 fractions of 5-8·5 Gy after a single application showed encouraging local control and survival outcomes with acceptable toxicities ^32^. Patients with Cervical cancer who contract COVID 19 infection with limited or mild symptoms could continue their Radiotherapy sessions ^33^. Dedicated Linear accelerators with staff donning Personal Protective Equipment (PPE) and patient access corridors in the hospital could facilitate such treatments.

The strengths of the study are its significant patient number with survival outcomes reported across both the groups. The median follow up duration was 37 months. Selection bias has been addressed by two investigators individually assessing each entry against the treatment records. Recall bias was minimised by reliance on hospital records rather than telephonic interviews. Telephonic interactions were only done to encourage the patient to visit the hospital where their follow up information could be verified. The potential drawbacks of this study include its retrospective nature and patient attrition (lost to follow up). About 95 patients were assessed for eligibility and removed from the final analysis as they were categorised as Registered only without access to any treatment or follow up records.

## Conclusion

In this retrospective cohort study, we find that delays and disruptions in cervical cancer care during the COVID-19 pandemic led to tumour upstaging and worsened survival outcomes. This highlights the urgent need for developing strategies aimed at optimal resource utilisation and minimizing delays in the event of a future pandemic.

## Data Availability

All data produced in the present work are contained in the manuscript

## Acknowledgements

None

## Prior Presentation and pre-prints

Part of results of this study were presented as a physical poster at ESMO Asia 2024 in Singapore. It is intended to submit a pre-print of this paper with medrxiv

## Availability of data and materials

The data underlying the results presented in this article may be accessible to researchers upon reasonable request made to the corresponding author

## Author contributions

1. Dr Venkata Krishna Vamsi Gade (FRCR) (Writing original draft – lead, Data curation – lead, Formal analysis-Lead, Methodology – supporting, Validation – supporting, Data Visualization - supporting)
2. Dr Jyoti Sharma (MD) (Data curation – supporting, Data Visualization – supporting, Validation – supporting)
3. Mr Sougata Maity (Msc) (Data curation – supporting, Formal analysis – supporting, Data Visualization – Lead)
4. Dr Tapesh Bhattacharyya (FRCR) (Data curation – supporting, Formal analysis-supporting, Methodology – supporting, Validation – supporting)
5. Dr Srinivasa GY (MD) (Data curation – supporting, Supervision – supporting)
6. Dr Dharmendra Kumar Sah (MD) (Data curation – supporting, Formal analysis – supporting. Data Visualization – supporting, Validation – supporting)
7. Dr Santam Chakraborty (MD) (Conceptualization – supporting, Supervision – supporting, Validation – Lead, Data Visualization – supporting, writing original draft – supporting)
8. Dr Bhavana Rai (MD) (Conceptualization – Lead, Supervision – Lead, Validation – supporting, Data Visualization – supporting, writing original draft – supporting)
9. Dr Sushmita Ghoshal (MD) (Conceptualization – supporting, Supervision – supporting, Validation – supporting)

All authors have reviewed and approved the final manuscript

## APPENDIX

**Appendix Table 1.**
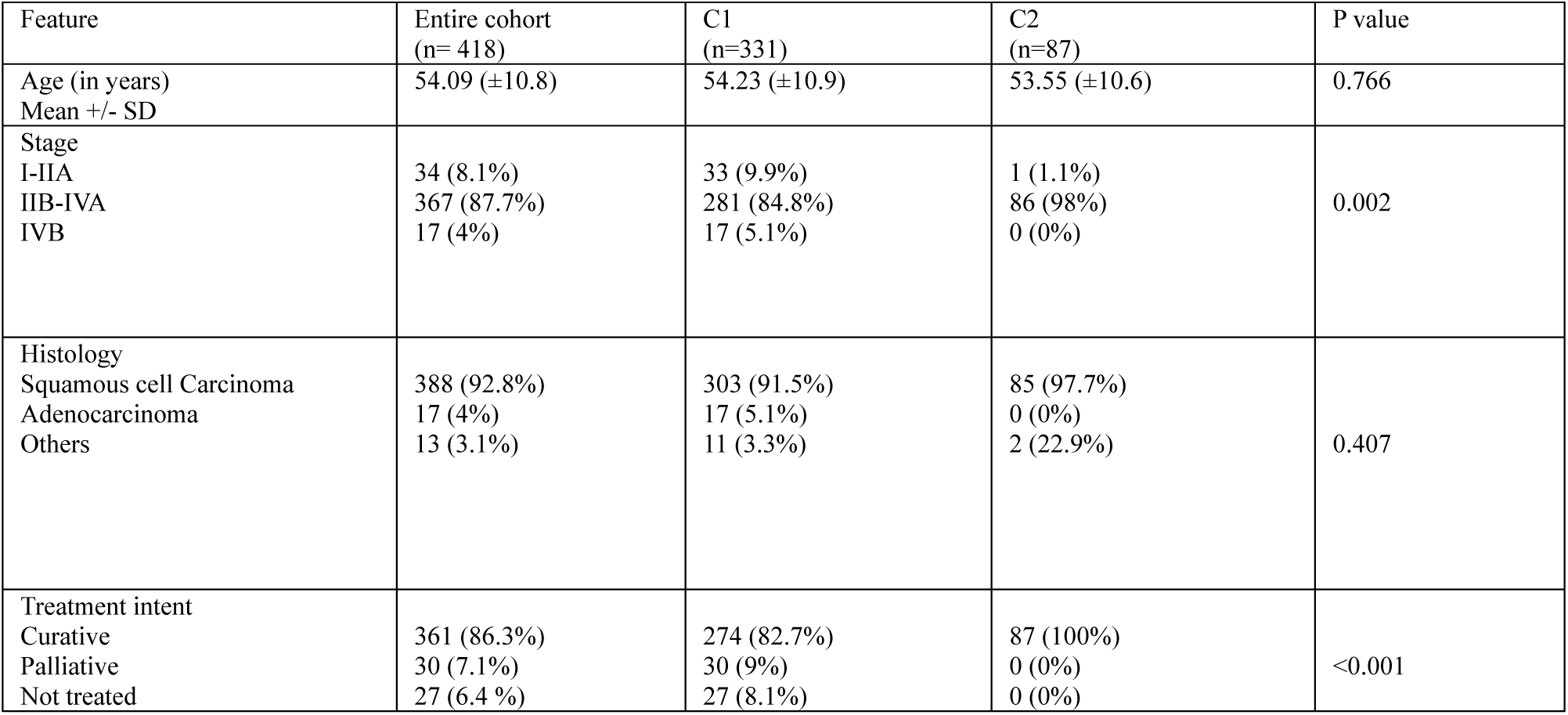
Baseline characteristics of the study cohort categorised centre wise.

| Feature | Entire cohort<br>(n= 418) | C1<br>(n=331) | C2<br>(n=87) | P value |
| --- | --- | --- | --- | --- |
| Age (in years)<br>Mean +/- SD | 54.09 (±10.8) | 54.23 (±10.9) | 53.55 (±10.6) | 0.766 |
| Stage<br>I-IIA<br>IIB-IVA<br>IVB | 34 (8.1%)<br>367 (87.7%)<br>17 (4%) | 33 (9.9%)<br>281 (84.8%)<br>17 (5.1%) | 1 (1.1%)<br>86 (98%)<br>0 (0%) | 0.002 |
| Histology<br>Squamous cell Carcinoma<br>Adenocarcinoma<br>Others | 388 (92.8%)<br>17 (4%)<br>13 (3.1%) | 303 (91.5%)<br>17 (5.1%)<br>11 (3.3%) | 85 (97.7%)<br>0 (0%)<br>2 (22.9%) | 0.407 |
| Treatment intent<br>Curative<br>Palliative<br>Not treated | 361 (86.3%)<br>30 (7.1%)<br>27 (6.4 %) | 274 (82.7%)<br>30 (9%)<br>27 (8.1%) | 87 (100%)<br>0 (0%)<br>0 (0%) | <0.001 |

**Appendix Table 2.** Treatment related details and patterns of spread categorised centre wise. (Note – Others in the treatment column includes those that received EBRT alone but no brachytherapy, surgery alone without adjuvant radiotherapy and those offered best supportive care).

| Feature | Entire cohort (n= 418) | C1<br>(n=331) | C2<br>(n=87) | P value |
| --- | --- | --- | --- | --- |
| Treatment details |  |  |  |  |
| Definitive RT followed by Brachytherapy | 267 (63.8%) | 182(55%) | 85 (97.7%) | <0.001 |
| Definitive RT followed by Supplementary boost | 48 (11.4%) | 46(13.8%) | 2 (2.3%) |  |
| Surgery followed by Adjuvant RT | 30 (7.1%) | 30 (9%) | 0 (0%) |  |
| Palliative RT | 12 (2.8%) | 12 (3.6%) | 0 (0%) |  |
| Palliative Chemotherapy | 2 (0.4%) | 2 (0.6%) | 0 (0%) |  |
| Not treated | 27 (6.4%) | 27 (8.1%) | 0 (0%) |  |
| Others | 32 (7.6%) | 32 (9.6%) | 0 (0%) |  |
| Progression or failure | 118 (28.2%) | 112 (33.8%) | 6 (6.8%) | <0.001 |
| Patterns of failure |  |  |  | p=0.02 |
| Local alone | 10 (2.3%) | 8 (2.4%) | 2 (2.3%) |  |
| Regional alone | 4 (0.9%) | 4 (1.2%) | 0 (0%) |  |
| Metastatic alone | 14 (3.3%) | 14 (4.2%) | 0 (0%) |  |
| Local and Regional | 50 (11.9%) | 48 (14.5%) | 2 (2.3%) |  |
| Local, Regional and Metastatic | 40 (9.5%) | 38 (11.4%) | 2 (2.3%) |  |

**Appendix Table 3.**
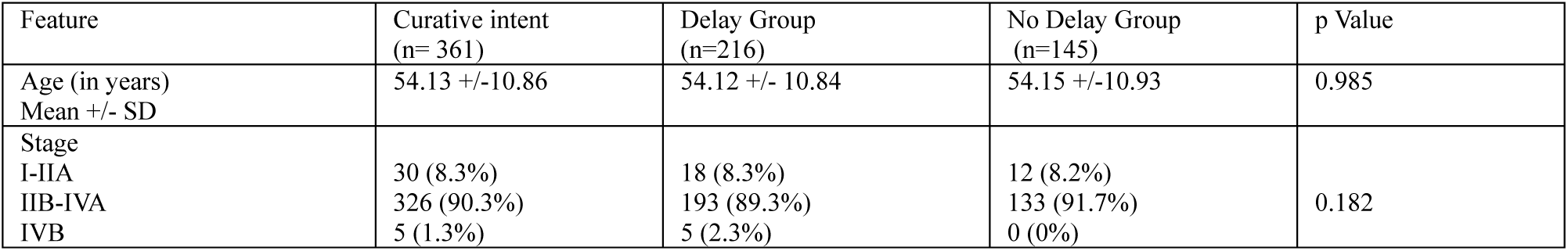
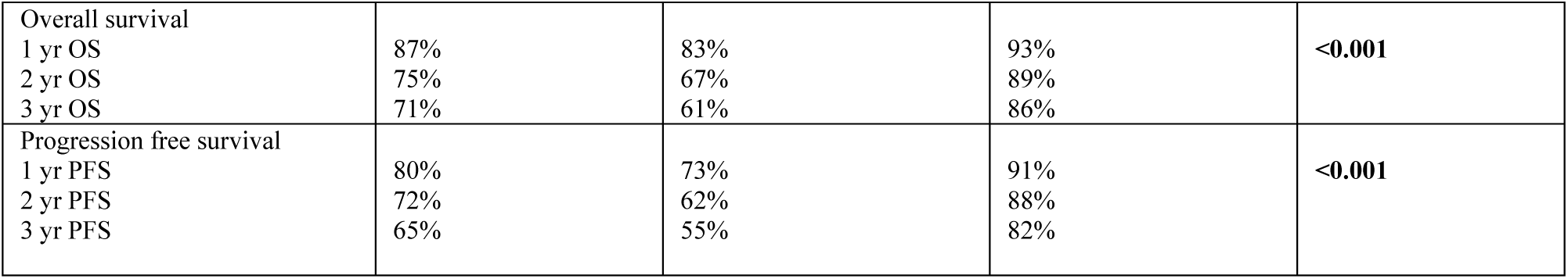
Features and survival outcomes of patients treated with curative intent.

**Appendix Figure 1.**
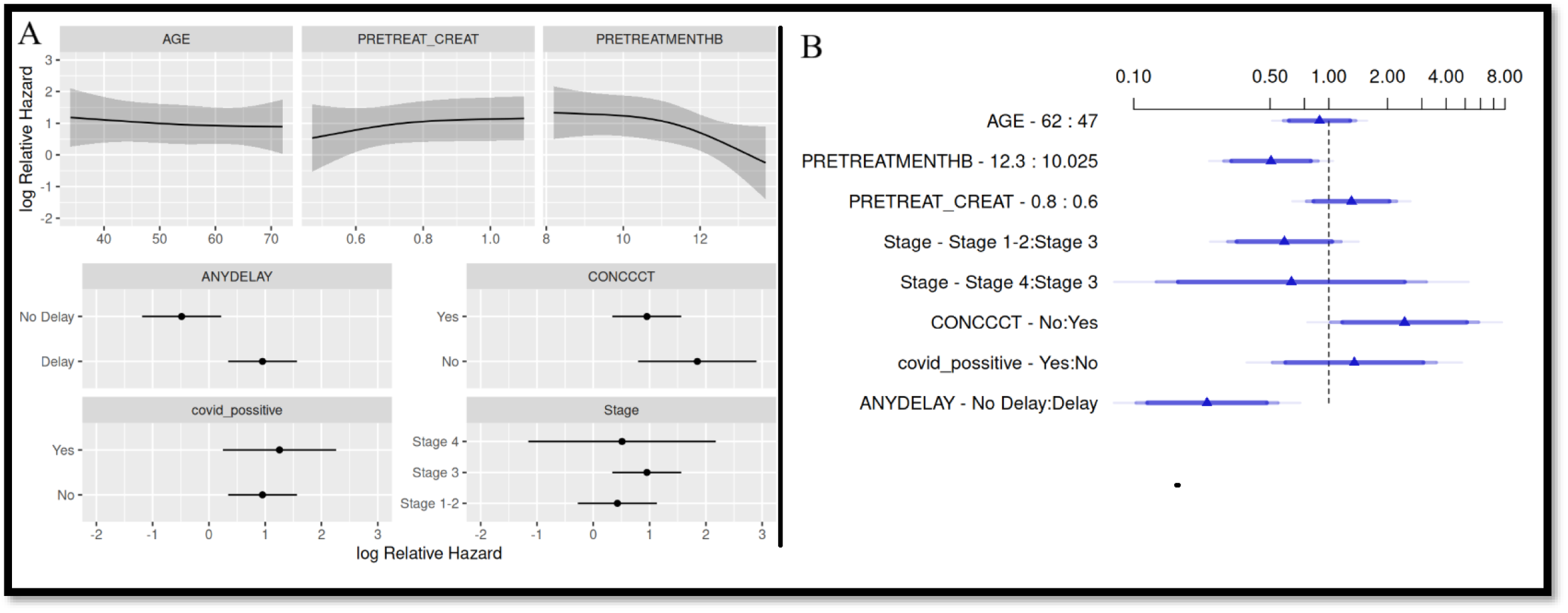
(A) - Log relative hazard plots for multivariate analysis of various factors affecting the OS in patients treated with curative chemoradiotherapy. (B)- Cox model Forest plot for multivariate analysis of various factors affecting the OS patients treated with curative chemoradiotherapy. Instead of studying delay as a numerical variable, this Multivariate analysis studies presence of delay as a categorical variable.

**Appendix Table 4.**
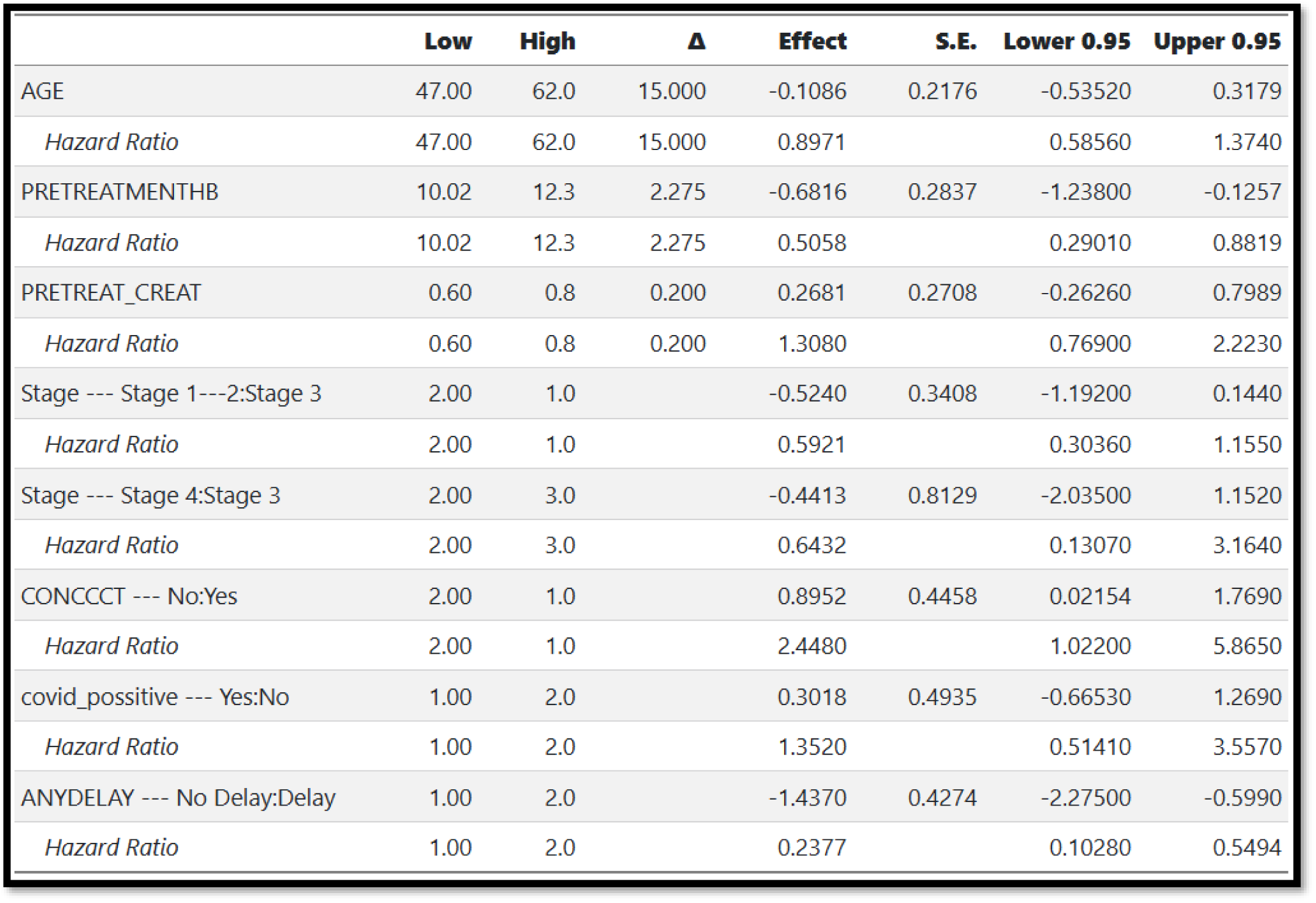
Multivariate analysis of various factors affecting the OS in patients treated with curative chemoradiotherapy. Instead of studying delay as a numerical variable, this Multivariate analysis studies presence of delay as a categorical variable.

**Appendix Figure 2.**
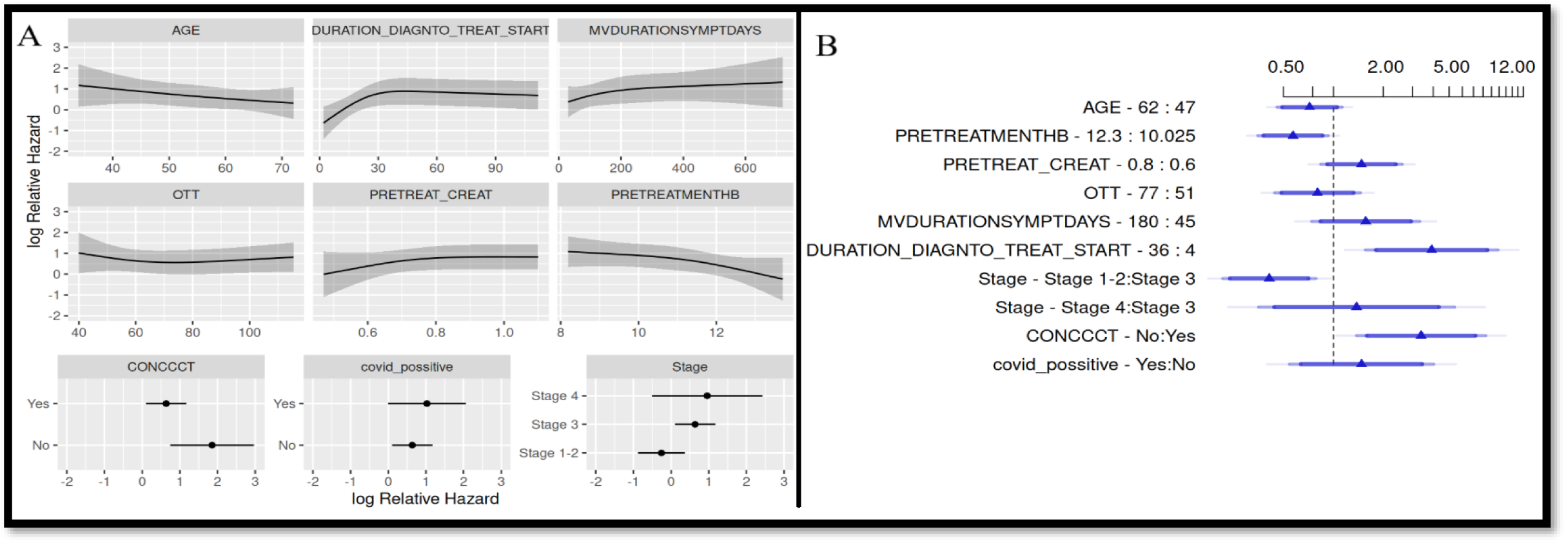
(A) - Log relative hazard plots for multivariate analysis of various factors affecting the PFS in patients treated with curative chemoradiotherapy. (B)-Cox model Forest plot for multivariate analysis of various factors affecting the PFS in patients treated with curative chemoradiotherapy. (OTT – Overall treatment time, MVDURATIONSYMPTOMDAYS – Duration of symptoms in days, CONCCT – Concurrent chemo given or not)

**Appendix Table 5.** Multivariate analysis of various factors affecting the PFS in patients treated with curative chemoradiotherapy. (OTT – Overall treatment time, MVDURATIONSYMPTOMDAYS – Duration of symptoms in days, CONCCT – Concurrent chemo given or not)

|  | **Low** | **High** | **Δ** | **Effect** | **S.E.** | **Lower**<br>**0.95** | **Upper 0.95** |
| --- | --- | --- | --- | --- | --- | --- | --- |
| AGE | 47.00 | 62.0 | 15.000 | -0.3302 | 0.2295 | -0.7801 | 0.11970 |
| *Hazard Ratio* | 47.00 | 62.0 | 15.000 | 0.7188 |  | 0.4584 | 1.12700 |
| PRETREATMENTHB | 10.02 | 12.3 | 2.275 | -0.5607 | 0.2483 | -1.0470 | -0.07398 |
| *Hazard Ratio* | 10.02 | 12.3 | 2.275 | 0.5708 |  | 0.3508 | 0.92870 |
| PRETREAT_CREAT | 0.60 | 0.8 | 0.200 | 0.3911 | 0.2874 | -0.1723 | 0.95450 |
| *Hazard Ratio* | 0.60 | 0.8 | 0.200 | 1.4790 |  | 0.8417 | 2.59700 |
| OTT | 51.00 | 77.0 | 26.000 | -0.2206 | 0.3029 | -0.8142 | 0.37310 |
| *Hazard Ratio* | 51.00 | 77.0 | 26.000 | 0.8021 |  | 0.4430 | 1.45200 |
| MVDURATIONSYMPTDAYS | 45.00 | 180.0 | 135.000 | 0.4502 | 0.3810 | -0.2965 | 1.19700 |
| *Hazard Ratio* | 45.00 | 180.0 | 135.000 | 1.5690 |  | 0.7434 | 3.31000 |
| DURATION_DIAGNTO_TREAT_START | 4.00 | 36.0 | 32.000 | 1.3660 | 0.4687 | 0.4478 | 2.28500 |
| *Hazard Ratio* | 4.00 | 36.0 | 32.000 | 3.9220 |  | 1.5650 | 9.82700 |
| Stage --- Stage 1---2:Stage 3 | 2.00 | 1.0 |  | -0.8912 | 0.3297 | -1.5370 | -0.24510 |
| *Hazard Ratio* | 2.00 | 1.0 |  | 0.4102 |  | 0.2150 | 0.78260 |
| Stage --- Stage 4:Stage 3 | 2.00 | 3.0 |  | 0.3220 | 0.6929 | -1.0360 | 1.68000 |
| *Hazard Ratio* | 2.00 | 3.0 |  | 1.3800 |  | 0.3548 | 5.36600 |
| CONCCCT --- No:Yes | 2.00 | 1.0 |  | 1.2200 | 0.4563 | 0.3258 | 2.11500 |
| *Hazard Ratio* | 2.00 | 1.0 |  | 3.3880 |  | 1.3850 | 8.28600 |
| covid_possitive --- Yes:No | 1.00 | 2.0 |  | 0.3912 | 0.5102 | -0.6088 | 1.39100 |
| *Hazard Ratio* | 1.00 | 2.0 |  | 1.4790 |  | 0.5440 | 4.01900 |

**Appendix Figure 3.**
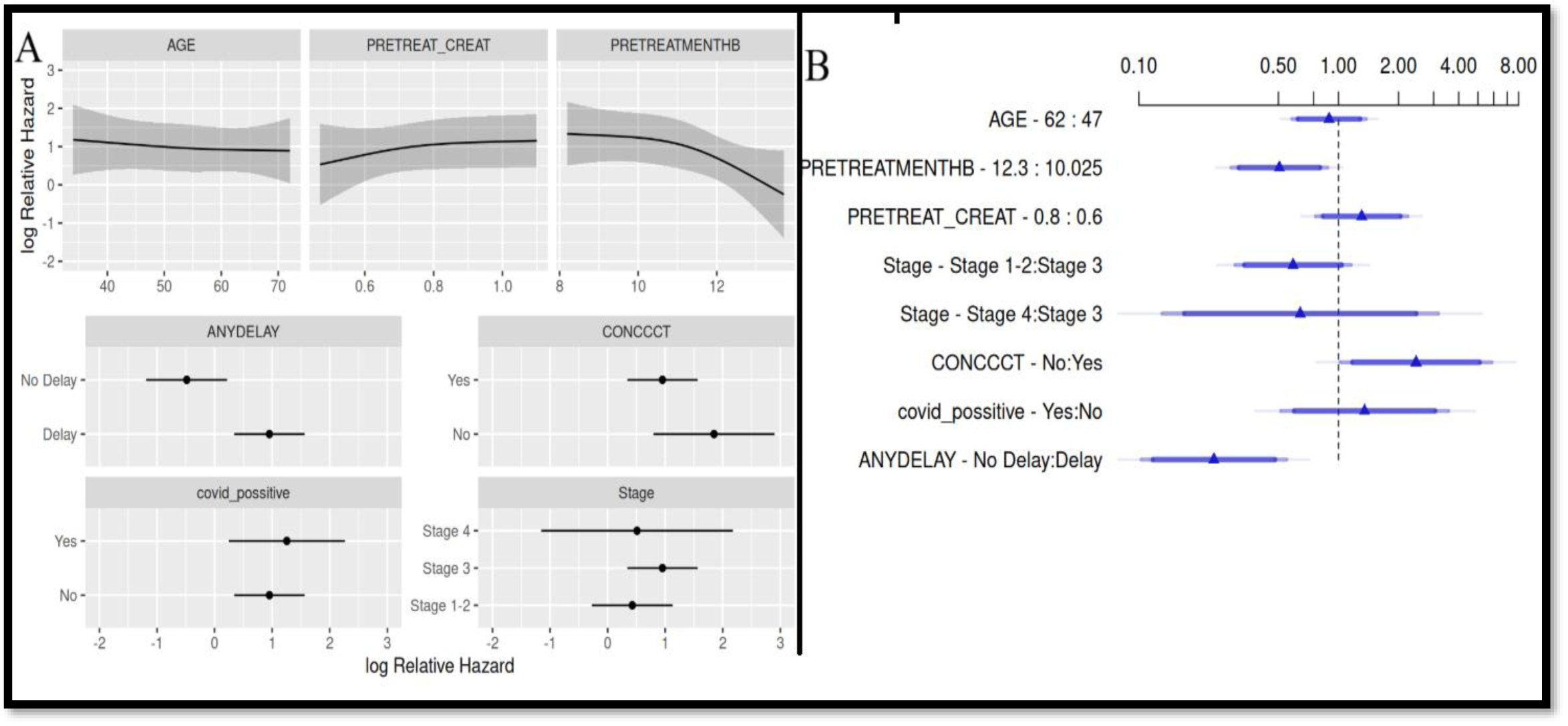
(A) - Log relative hazard plots for multivariate analysis of various factors affecting the PFS in patients treated with curative chemoradiotherapy. (B)-Cox model Forest plot for multivariate analysis of various factors affecting the PFS in patients treated with curative chemoradiotherapy. Instead of studying delay as a numerical variable, this Multivariate analysis studies presence of delay as a categorical variable.

**Appendix Table 6.**
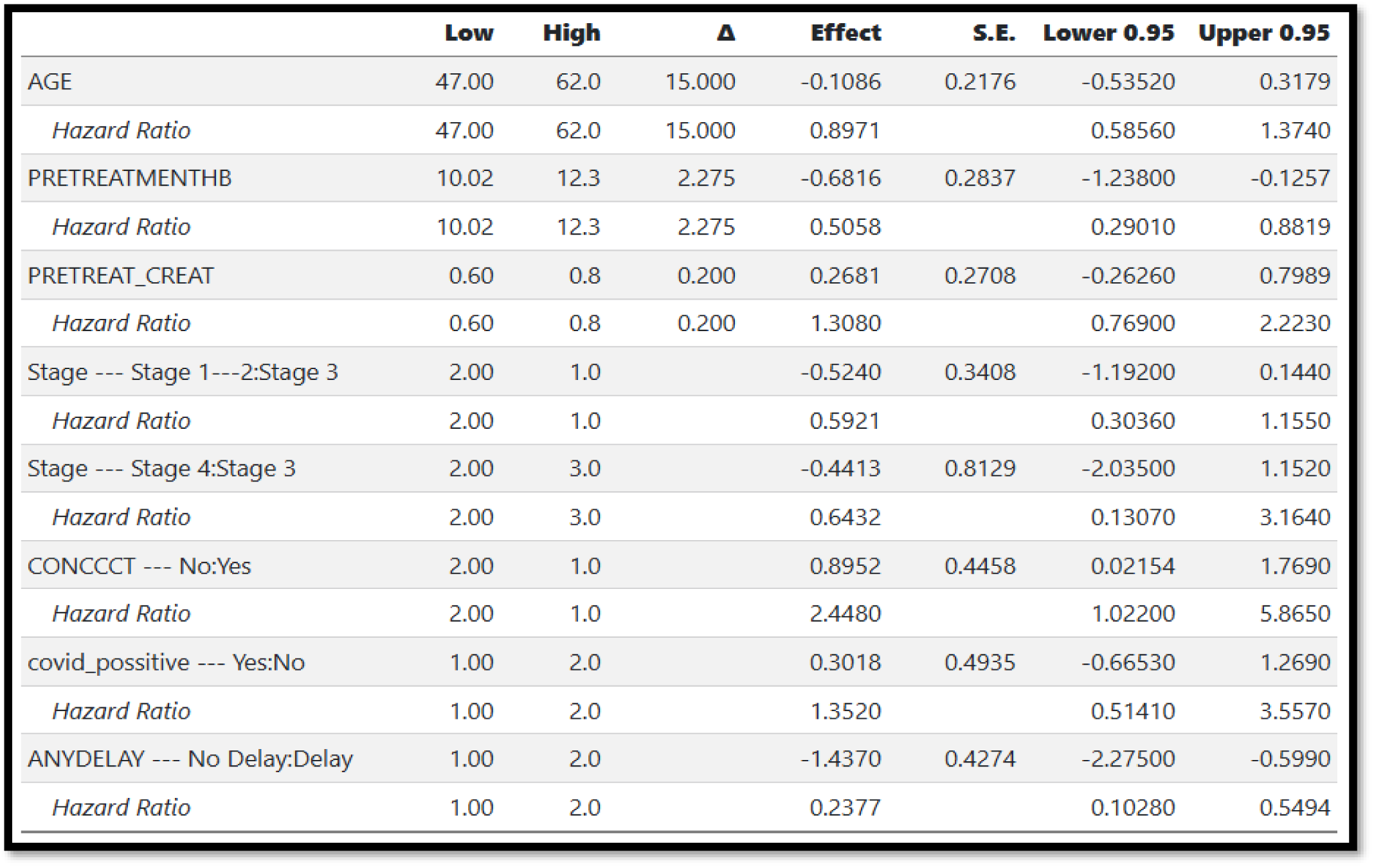
Multivariate analysis of various factors affecting the PFS in patients treated with curative chemoradiotherapy. Instead of studying delay as a numerical variable, this Multivariate analysis studies presence of delay as a categorical variable.

**Appendix Figure 4.**
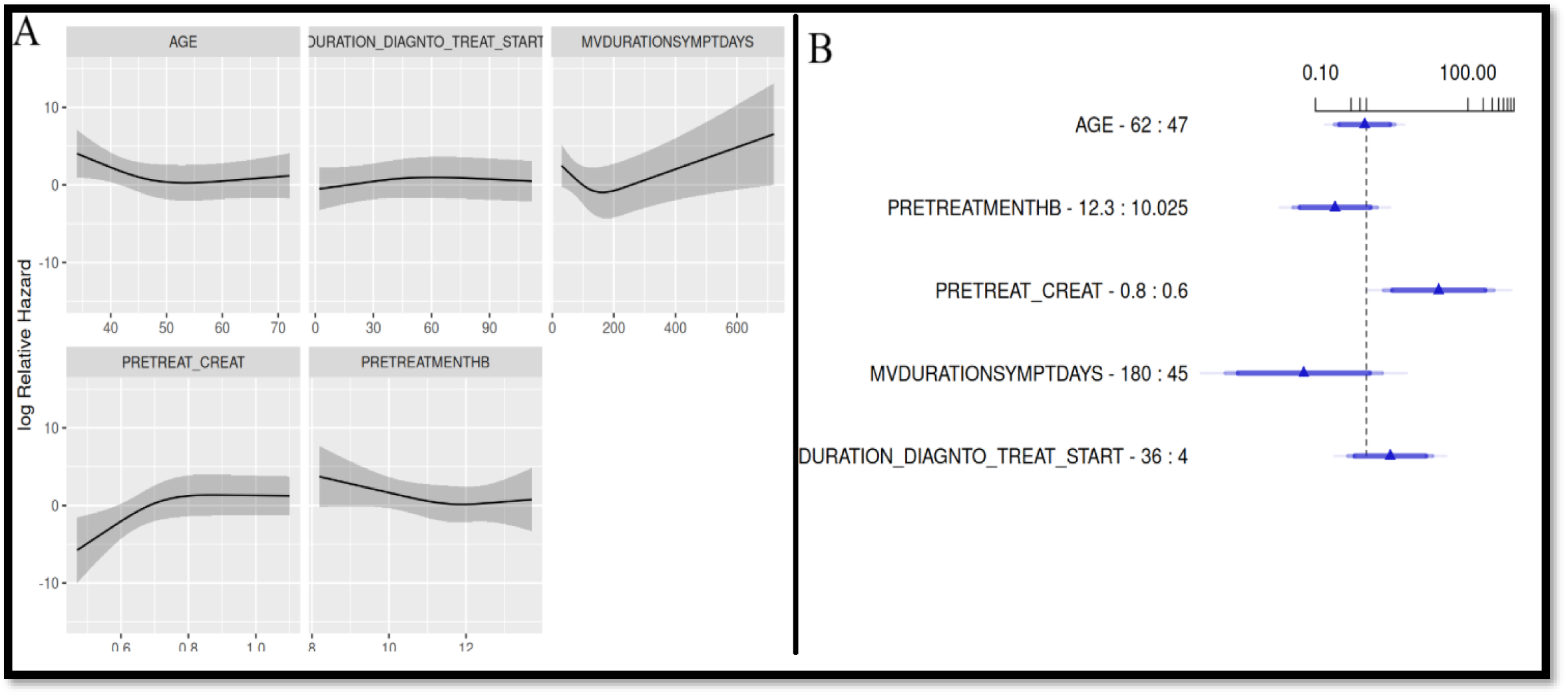
(A) - Log relative hazard plots for multivariate analysis of various factors affecting the OS in patients treated with Palliative intent. (B)-Cox model Forest plot for multivariate analysis of various factors affecting the OS in patients treated with Palliative intent.

**Appendix Table 7.** Multivariate analysis of various factors affecting the OS in patients treated with Palliative intent.

|  | **Low** | **High** | **Δ** | **Effect** | **S.E.** | **Lower 0.95** | **Upper 0.95** |
| --- | --- | --- | --- | --- | --- | --- | --- |
| AGE | 47.00 | 62.0 | 15.000 | -0.07326 | 0.6972 | -1.440000 | 1.2930 |
| *Hazard Ratio* | 47.00 | 62.0 | 15.000 | 0.92940 |  | 0.237000 | 3.6440 |
| PRETREATMENTHB | 10.02 | 12.3 | 2.275 | -1.41500 | 0.9729 | -3.322000 | 0.4921 |
| *Hazard Ratio* | 10.02 | 12.3 | 2.275 | 0.24300 |  | 0.036090 | 1.6360 |
| PRETREAT_CREAT | 0.60 | 0.8 | 0.200 | 3.28300 | 1.2760 | 0.781200 | 5.7840 |
| *Hazard Ratio* | 0.60 | 0.8 | 0.200 | 26.65000 |  | 2.184000 | 325.1000 |
| MVDURATIONSYMPTDAYS | 45.00 | 180.0 | 135.000 | -2.83000 | 1.8150 | -6.388000 | 0.7271 |
| *Hazard Ratio* | 45.00 | 180.0 | 135.000 | 0.05899 |  | 0.001682 | 2.0690 |
| DURATION_DIAGNTO_TREAT_START | 4.00 | 36.0 | 32.000 | 1.08700 | 0.9820 | -0.837700 | 3.0110 |
| *Hazard Ratio* | 4.00 | 36.0 | 32.000 | 2.96500 |  | 0.432700 | 20.3200 |

